# Infant EEG profiles prospectively differentiate temperament and early mental health risk in childhood

**DOI:** 10.64898/2026.06.15.26355713

**Authors:** Dashiell D. Sacks, Owen Forbes, Charles A. Nelson, Michelle Bosquet Enlow

## Abstract

**Background:** EEG provides a scalable method for elucidating neurophysiological characteristics that may distinguish mental health risk early in life, when symptoms are often non-specific, transdiagnostic, and pluripotential. Most prior studies have examined cross-sectional associations between individual EEG metrics and singular outcomes, potentially overlooking integrated patterns of neurophysiological organization. We applied data-driven clustering to infant baseline EEG to derive neurophysiological profiles and examined whether these profiles prospectively differentiated temperament and psychopathology domains in childhood.

**Methods:** Participants were (*N* = 360; 46% female) from a longitudinal community cohort followed from infancy to age 7 years. Baseline EEG was collected in infancy (*M*age = 7.81 months). Neurophysiological profiles were derived from spectral features (band-limited periodic power, peak frequency characteristics, and aperiodic exponent) using Bayesian model averaging of multiple clustering algorithms. Bayesian mixed-effects models tested profile differences in parent-reported temperament (surgency, negative affectivity, regulation/effortful control) across infancy and ages 3, 5, and 7 years, and child internalizing and externalizing symptoms at 5 and 7 years.

**Results:** Consensus clustering identified four infant neurophysiological profiles characterized by: (1) elevated alpha/beta power, (2) low-frequency-dominant power, (3) globally attenuated oscillatory power, and (4) faster frequency-shifted dynamics. The profiles showed graded differentiation across childhood in effortful control (Cluster 1>2>3>4), with strong evidence for higher effortful control in Clusters 1/2 relative to Clusters 3/4 (posterior probabilities > .95). Additional differentiation was observed across surgency, negative affectivity, and psychopathology symptoms. Clusters 3/4 showed higher internalizing and externalizing symptom probabilities relative to Clusters 1/2, particularly Cluster 2, which also showed lower surgency relative to Clusters 1/3/4.

**Conclusions:** Infant EEG-derived neurophysiological profiles prospectively differentiated temperament and psychopathology outcomes in childhood. With ongoing research, data-driven EEG profiling may provide a scalable, biologically informed framework for early mental health risk stratification prior to the consolidation of stable psychiatric diagnoses.

---

Mental health disorders impose an immense global burden, markedly reducing quality of life and functioning among those affected and resulting in estimated annual economic losses of over USD 5 trillion (Arias et al., 2022). More than half of all mental disorders have their onset in childhood and adolescence, and psychopathology can be identified as early as toddlerhood (Bufferd et al., 2011; Egger & Angold, 2006; Kessler et al., 2005). Earlier onset is associated with poorer long-term outcomes, including greater functional impairment, chronicity, and reduced treatment responsivity (Copeland et al., 2015; Vergunst et al., 2023). However, in youth, the identification and classification of mental health disorders remains challenging, as psychopathology symptoms are often non-specific, transdiagnostic, and pluripotential (Conway et al., 2022; Sawrikar et al., 2022). Thus, there is a need for biologically informed approaches that can support early risk stratification prior to the consolidation of more stable psychiatric diagnoses.

Electroencephalography (EEG) is a particularly appealing modality for investigating early neurodevelopmental risk, as it is portable (and thus scalable), relatively affordable, non-invasive, and generally well tolerated in pediatric populations (Nelson et al., 2026). EEG has been used extensively to examine individual differences in neurophysiological development relevant to cognition and mental health, including in child psychiatry research (see e.g., (McVoy et al., 2019). However, much of the existing literature has focused on testing associations between individual EEG measures (e.g., power in frequency bands or derivatives such as the theta/beta ratio or frontal alpha asymmetry) and specific diagnoses or characteristics. Although this research has yielded important insights, such approaches may be limited in their ability to capture the heterogeneous and dynamic nature of early neurodevelopment and emergent psychopathology. In this context, data-driven clustering offers a useful framework for integrating multiple EEG features to identify patterns of neurophysiological organization, yielding biologically based profiles – potential neurodevelopmental endophenotypes – that may prospectively differentiate risk for later emotional, behavioral and mental health outcomes.

To this end, Forbes et al. (2022) applied a novel unsupervised clustering pipeline to resting-state EEG in an adolescent sample (*N* = 59; preliminary cross-sectional study) and compared the derived neurophysiological profiles across measures of mental health and wellbeing, sleep, and cognitive functioning. More specifically, various summary EEG spectral features (frequency band power, individual alpha frequency, spectral edge frequency, entropy) were clustered using a consensus approach that integrated solutions across multiple common clustering algorithms, yielding five neurophysiological profiles. These profiles differed across levels of self-reported psychological distress, wellbeing, sleep quality, and various cognitive domains cross-sectionally. This approach has distinct strengths, as neurophysiological profiles were derived from integrated EEG measures independent of symptom or diagnostic information, thereby reflecting biologically driven differences in neural organization that distinguished concurrent measures of mental health, sleep, and cognitive outcomes.

Despite the promise of this work, important questions remain about whether similar neurophysiological profiles can be identified earlier in childhood and whether such profiles can prospectively differentiate psychopathology risk. Given that psychopathology symptoms can emerge from early childhood but are often difficult to characterize during this period, such approaches may be particularly valuable early in development. The current study applied a data-driven approach to identify biologically defined baseline EEG profiles in a large infant sample (*N* = 360), thereby applying this approach to one of the earliest developmental periods in which neurophysiological organization can be meaningfully characterized. Specifically, we incorporated child-oriented EEG preprocessing and contemporary spectral parameterization to derive key summary features and applied a Bayesian model averaging framework for clustering to identify early emerging neurophysiological profiles in infancy. We then examined whether these EEG-derived profiles prospectively differentiated behavioral and mental health measures assessed longitudinally across childhood. Given the developmental stage of the sample and the evolving nature of psychopathology during this period, we focused on two primary parent-reported outcome domains: (a) internalizing and externalizing symptoms assessed at ages 5 and 7 years, and (b) temperament traits – early-emerging individual differences in reactivity and self-regulation that represent established transdiagnostic risk factors for psychopathology (Sacks et al., 2026) – assessed in infancy and at ages 3, 5, and 7 years.

## Methods

### Participants

Participants were part of a longitudinal study designed to examine early neural correlates of emotion processing during development, beginning in infancy. Families were recruited from a registry of local births comprising families who had indicated willingness to participate in developmental research. By design, families were enrolled in the parent study when the children were 5, 7, or 12 months old (*M*age months = 7.81, *SD* = 2.82, 46% Female). Exclusion criteria included known prenatal or perinatal complications, maternal use of medications during pregnancy that may significantly impact fetal brain development (i.e., anticonvulsants, antipsychotics, opioids), pre- or post-term birth (≥ ±3 weeks from due date), developmental delay, uncorrected vision difficulties, and neurological disorder or trauma. After enrollment, families’ data were excluded from analyses if the child was diagnosed with an autism spectrum disorder or a genetic or other condition known to influence neurodevelopment. *N* = 360 participants were retained after EEG preprocessing and included in the infant EEG clustering analysis.

### Data Collection

The Institutional Review Board at Boston Children’s Hospital approved all study methods and procedures, and parents provided written informed consent prior to the initiation of study activities. Caregivers completed questionnaires via an online survey at each time point (infancy, 3 years, 5 years, 7 years). Questionnaires relevant to the current analyses included assessments of sociodemographic characteristics (collected at infancy visit), child temperament (collected at all time points), and child psychopathology symptoms (collected at 5 years and 7 years). EEG data were collected in infancy during an in-person study visit.

### Measures

#### Sociodemographic Characteristics

Sociodemographic characteristics collected included child age, sex assigned at birth (hereafter “sex”), ethnicity, race, parental educational attainment, and annual household income.

#### Child Temperament

Temperament was measured at all timepoints via age-appropriate parent-report questionnaires that share a common three-factor structure: negative affectivity, surgency/extraversion, and effortful control (orienting/regulation in infancy). Negative affectivity represents the tendency to experience negative emotions, such as anger, fear, frustration, and sadness; surgency represents activity level, approach tendencies, impulsivity, and high-intensity positive affect; effortful control represents voluntary regulation of attention, emotion, and behavior (Rothbart & Bates, 2006). In infancy, parents completed the 91-item Infant Behavior Questionnaire – Revised, Short Form (IBQ-R-SF; Putnam et al., 2014), Cronbach’s α values in the current sample were α = .86 for negative affectivity, α = .92 for surgency, and α = .81 for orienting/regulation. At age 3 years, parents completed the 107-item Early Childhood Behavior Questionnaire – Short Form (ECBQ-SF; Putnam et al., 2010). Cronbach’s α values were: negative affectivity α = .85, surgency α = .82, effortful control α = .84. At ages 5 and 7 years, parents completed the 36-item Children’s Behavior Questionnaire – Very Short Form (CBQ-VSF; Putnam & Rothbart, 2006). Cronbach’s α values were: negative affectivity α = .70, surgency α = .74, effortful control α = .72 at 5 years; negative affectivity α = .74, surgency α = .75, effortful control α = .75 at 7 years. For each scale, items were rated on a 7-point scale (1 = never, 7 = always); item scores were averaged to create scale scores, with higher scores indicating greater levels of that temperament dimension.

#### Child Mental Health

To measure child psychopathology symptoms across internalizing and externalizing domains, the Child Behavior Checklist (CBCL) for ages 1.5 to 5 years was administered at age 5 years, and the CBCL for ages 6 to 18 years at age 7 years (Achenbach & Rescorla, 2000; Achenbach et al., 2001; Achenbach & Edelbrock, 1991; Achenbach & Rescorla, 2014). Parents rated each questionnaire item on a 3-point scale ranging from 0 (“Not True/Rarely”) to 2 (“Very True/Often”). Item scores were summed to calculate raw scores. Subscale raw scores were age- and sex-normed, with normed scores expressed as the standard T-score metric (population mean of 50 and a standard deviation of 10). The CBCL composite score for internalizing symptoms comprises the following syndrome scales at age 5 years: emotionally reactive, anxious/depressed, somatic complaints, and withdrawn. At age 7 years, internalizing symptoms comprises anxious/depressed, somatic complaints, and withdrawn/depressed. The CBCL composite scores for externalizing problems comprise attention problems and aggressive behavior at age 5 years, and aggressive behavior and rule-breaking behavior at age 7 years. For internalizing symptoms, Cronbach’s α in this sample was .82 at 5 years and .82 at 7 years. For externalizing symptoms, Cronbach’s α in this sample was .90 at 5 years and .89 at 7 years.

### EEG

#### Acquisition

Continuous scalp EEG was recorded using a 128-channel HydroCel Geodesic Sensor Net (HGSN; Electrical Geodesics, Inc.) during the infancy assessment. The four electrooculogram channels were removed from the net for infant comfort. The net was connected to a NetAmps 300 amplifier (Electrical Geodesics, Inc.) and referenced online to a single vertex electrode (Cz). Data were sampled at 500 Hz. Channel impedances were kept at or below 100 kΩ, which is within recommended guidelines given the high-input impedance capabilities of this system’s amplifier. Infants sat on their caregiver’s lap in a dim room and watched a computer-generated video of moving toys while baseline EEG was recorded for 2 minutes.

#### Preprocessing

The EEG data were preprocessed in a manner consistent with recent studies in this and related cohorts (Sacks, Levin, et al., 2025; Sacks, Valdes, et al., 2025); Wilkinson et al. (2024). Raw Netstation (Electrical Geodesics, Inc) EEG files were exported into the MATLAB MAT-file format for preprocessing in MATLAB (version R2023a) using the Batch Automated Processing Platform (BEAPP; Levin et al., 2018), with integrated Harvard Automated Preprocessing Pipeline for EEG (HAPPE; Gabard-Durnam et al., 2018). Data were high-pass filtered at a 1Hz and low-pass filtered at 100Hz. Data were resampled to 250Hz and then preprocessed using the HAPPE module, which includes line noise removal using CleanLine multi-taper regression, bad channel rejection through evaluation of the normed joint probability of the average log power, and artifact removal using combined wavelet-enhanced independent component analysis (ICA) and Multiple Artifact Rejection Algorithm (MARA; Winkler et al., 2015). In addition to the 10-20 electrodes, the following channels were used for MARA: 28, 19, 4, 117, 13, 112, 41, 47, 37, 55, 87, 103, 98, 65, 67, 77, 90, 75. After artifact removal, bad channels were interpolated using spherical interpolation. Data were rereferenced to the average reference, detrended to the signal mean, and segmented into 2-second epochs. HAPPE’s amplitude and joint probability criteria were used to reject epochs contaminated with artifact. Recordings with fewer than 20 segments (i.e., at least 40 seconds of total EEG), percent good channels < 80%, percent independent components rejected >80%, mean artifact probability of components kept > 0.3, and/or percent variance retained < 25% were rejected (*N* = 45, 11%).

#### Power spectral density and spectral parameterization

Power spectral density was calculated in the BEAPP Power Spectral Density (PSD) module using a multitaper spectral analysis with three orthogonal tapers. For each electrode, the PSD was averaged across segments, and then further averaged across all channels (10-20 and MARA; ‘33’, ‘22’, ‘9’, ‘122’, ‘28’, ‘24’, ‘19’, ‘11’, ‘4’, ‘124’, ‘117’, ‘13’, ‘112’, ‘45’, ‘41’, ‘36’, ‘37’, ‘55’, ‘87’, ‘104’, ‘103’, ‘108’, ‘47’, ‘52’, ‘67’, ‘62’,’77’, ‘92’, ‘98’, ‘58’, ‘65’, ‘70’, ‘75’, ‘83’, ‘90’, ‘96’). The PSD was parameterized using a modified version of SpecParam (Donoghue et al., 2020) applied in recent studies of infants (Chung et al., 2025; Sacks, Levin, et al., 2025; Wilkinson et al., 2024). In this version, the robust_ap_fit function is modified, so that the initial estimate of the flattened power spectra (flatspec) has a baseline elevated such that the lowest point is ≥ 0. The SpecParam model was fit across a 2–55 Hz frequency range, in the fixed mode (no spectral knee), with peak_width_limits set to [0.5, 18.0], max_n_peaks = 7, and peak_threshold = 2. Mean R^2^ for the full sample was 0.996 (*SD* = 0.013). Mean estimated error for the full sample was 0.013 (*SD =* 0.013). The following summary measures were extracted: Periodic (oscillatory) power was extracted by integrating spectral power after subtraction of the modeled aperiodic component within the canonical frequency ranges of theta (4-6Hz), low alpha (6-9Hz), high alpha (9-12Hz), low beta (12-20Hz), and high beta (20-30Hz). Peak frequencies within the theta/alpha range (4-12Hz) and high beta range were derived by extracting maxima from the periodic spectrum after subtraction of the modeled aperiodic component. Finally, we extracted the aperiodic exponent (a positive value; exponent x from the 1/f^x^ distribution, with a larger exponent value indicating a steeper slope).

### Data Analysis

#### Unsupervised clustering of EEG features with Bayesian model averaging

Statistical analyses were conducted in R 4.3.2 (R Core Team, 2025). Periodic theta, low alpha, high alpha, low beta, and high beta power, alpha and beta peak frequencies, and the aperiodic exponent were preselected as clustering features. Because infants completed EEG data collection at ages 5, 7, or 12 months, we standardized features within these ages prior to clustering to minimize developmental confounding (i.e., excessive age loading of clusters), while preserving relative individual differences. To reduce dimensionality and mitigate collinearity among EEG features prior to clustering, principal component analysis (PCA) was applied to the standardized feature matrix using the prcomp() function from the base R *stats* package. Number of principal components was selected via inspection of the scree plot and cumulative proportion of variance explained. We implemented three complementary unsupervised clustering algorithms: k-means (kmeans() with 25 random starts), hierarchical clustering (hclust() with method “ward.D2”), and Gaussian mixture modeling (GMM; GMM() function from *ClusterR 1.3.5* (Mouselimis, 2020)). Cluster solutions (k = 3-10) were evaluated separately for each algorithm via assessment of multiple internal validation indices computed using *clusterCrit* 1.3.0 (Desgraupes, 2023): Calinski-Harabasz index, silhouette width, Dunn index, Davies-Bouldin index, Xie-Beni index, as well as Bayesian Information Criterion (BIC) for GMM. Cluster assignments from the three clustering approaches were then integrated using Bayesian model averaging (*ClusterBMA 0.2.0*; Forbes et al., 2023). This approach probabilistically integrates clustering solutions across algorithms to estimate consensus clusters, reducing dependence on any single clustering method.

#### Analysis of Cluster Differences in Temperament and Mental Health

To examine whether infant EEG-derived clusters differentiated temperament traits and psychopathology symptoms through age 7 years, we fit Bayesian mixed-effects models using *brms 2.23.0 (Bürkner, 2017)*. Cluster allocation was the main categorical predictor, and all models included sex as a covariate and a participant-level random intercept to account for repeated measures. For each outcome (surgency, negative affectivity, effortful control, internalizing, externalizing), three models were evaluated using the widely applicable information criterion (WAIC): (1) a cluster-only model estimating average cluster differences across ages, (2) an additive model including both cluster and age effects, and (3) a cluster * age interaction model allowing cluster differences to vary across developmental ages. Temperament traits were modeled across infancy and ages 3, 5, and 7 years; psychopathology symptoms were modeled across ages 5 and 7 years. For temperament outcomes, regression coefficients were assigned normal priors centered at zero with standard deviation 1, and intercepts were assigned normal priors centered at the midpoint of the scale (mean = 4, SD = 1). For internalizing and externalizing T-scores, regression coefficients were assigned normal priors (mean = 0, SD = 10), and intercepts were assigned normal priors centered on the expected population mean (mean = 50, SD = 10). Residual and random-effect standard deviations were assigned exponential(1) priors for temperament outcomes and exponential(0.1) priors for T-score outcomes. Prior sensitivity analyses were conducted by refitting all models with alternate prior specifications (more regularizing, more diffuse, and heavy-tailed Student-t priors) and comparing estimates. Cluster differences were evaluated using posterior contrasts between estimated marginal means computed with *emmeans 2.0.0 (Lenth & Piaskowski, 2025)*. For each pairwise cluster comparison, posterior samples were used to estimate directional posterior probabilities representing the probability that the mean value of the outcome measure was greater in one cluster than in another. Effect sizes (Cohen’s d) and 95% credible intervals were additionally reported.

### Results

Sample sociodemographic characteristics are presented in Table 1. Children were predominantly non-Hispanic White, and parental education and annual household income indicated middle to high socioeconomic status.

**Table 1.** Sample sociodemographic characteristics, collected at infancy (*N*=360).

| Variables | *n* | % |
| --- | --- | --- |
| Child sex |  |  |
| Male | 197 | 54.7 |
| Female | 163 | 45.3 |
| Child race |  |  |
| White | 287 | 79.7 |
| Black/African American | 7 | 1.9 |
| Asian | 18 | 5.0 |
| More than one race | 44 | 12.2 |
| Not reported | 4 | 1.1 |
| Child ethnicity |  |  |
| Non-Latino/a, Spanish, or Hispanic | 316 | 87.8 |
| Latino/a, Spanish, or Hispanic | 40 | 11.1 |
| Not reported | 4 | 1.1 |
| Maternal educational attainment |  |  |
| Less than high school | 2 | 0.6 |
| High school/GED | 13 | 3.6 |
| Associate degree | 8 | 2.2 |
| Bachelor's degree | 108 | 30.0 |
| Master's degree | 153 | 42.5 |
| M.D., Ph.D., J.D., or equivalent | 74 | 20.6 |
| Not reported | 2 | 0.6 |
| Paternal educational attainment |  |  |
| Less than high school | 1 | 0.3 |
| High school/GED | 24 | 6.7 |
| Associate degree | 18 | 5.0 |
| Bachelor's degree | 102 | 28.3 |
| Master's degree | 113 | 31.4 |
| M.D., Ph.D., J.D., or equivalent | 96 | 26.7 |
| Not reported | 6 | 1.7 |
| Annual family household income |  |  |
| Less than \$35,000 | 10 | 2.8 |
| \$35,000-\$49,000 | 16 | 4.4 |
| \$50,000-\$74,999 | 35 | 9.7 |
| \$75,000-\$99,000 | 52 | 14.4 |
| \$100,000 or more | 216 | 60.0 |
| Not reported | 31 | 8.6 |

### PCA

Based on visual inspection of the scree plot (Figure S1; Table S1) and application of the Kaiser criterion (eigenvalue > 1), the first four principal components were retained, which together explained 80.3% of the total variance (PC1: 29.4%, PC2: 19.4%, PC3: 18.5%, PC4: 12.9%). Alpha and beta power were dominant features in PC1, theta and alpha power and peak frequency in PC2 (19.4%), high beta power and alpha/beta peak frequency in PC3 (18.5%), and the aperiodic exponent in PC4 (12.9%) (Table 2).

**Table 2.** Component coefficients for PCA of frequency features.

| Feature | PC1 | PC2 | PC3 | PC4 |
| --- | --- | --- | --- | --- |
| Periodic Theta Power | -0.125 | **-0.738** | 0.160 | 0.068 |
| Periodic Low Alpha Power | **-0.457** | **-0.435** | -0.181 | -0.034 |
| Periodic High Alpha Power | **-0.543** | -0.002 | -0.254 | 0.134 |
| Periodic Low Beta Power | **-0.468** | 0.256 | 0.276 | 0.162 |
| Periodic High Beta Power | -0.363 | 0.061 | **0.495** | -0.153 |
| Alpha Peak Frequency | -0.346 | **0.400** | **-0.442** | -0.085 |
| Beta Peak Frequency | 0.086 | -0.178 | **-0.587** | 0.189 |
| Aperiodic Exponent (Slope) | 0.043 | -0.076 | -0.129 | **-0.940** |
| Total Proportion of Variance Explained | 0.294 | 0.194 | 0.185 | 0.129 |

Note. Coefficients with magnitude $\geq 0.4$ are in bold.

### Unsupervised Clustering

Unsupervised clustering was performed on the four principal components. Based on the internal validation indices (Calinski-Harabasz index, silhouette width, Dunn index, Davies-Bouldin index, Xie-Beni index, Bayesian Information Criterion (BIC) for GMM), we selected k = 4 for k-means, k = 7 for hierarchical clustering, and k = 3 for Gaussian mixture modeling (Figures S2-S4). The number of consensus clusters was determined by ClusterBMA’s regularization procedure, which pruned redundant components from an initial solution space spanning up to k = 7 clusters, yielding a final four-cluster solution (Cluster 1, *N* = 46; Cluster 2, *N* = 125; Cluster 3, *N* = 140; Cluster 4, *N* = 49). Mean absolute and periodic spectra in each cluster are shown in Figure 1.

**Figure 1.**
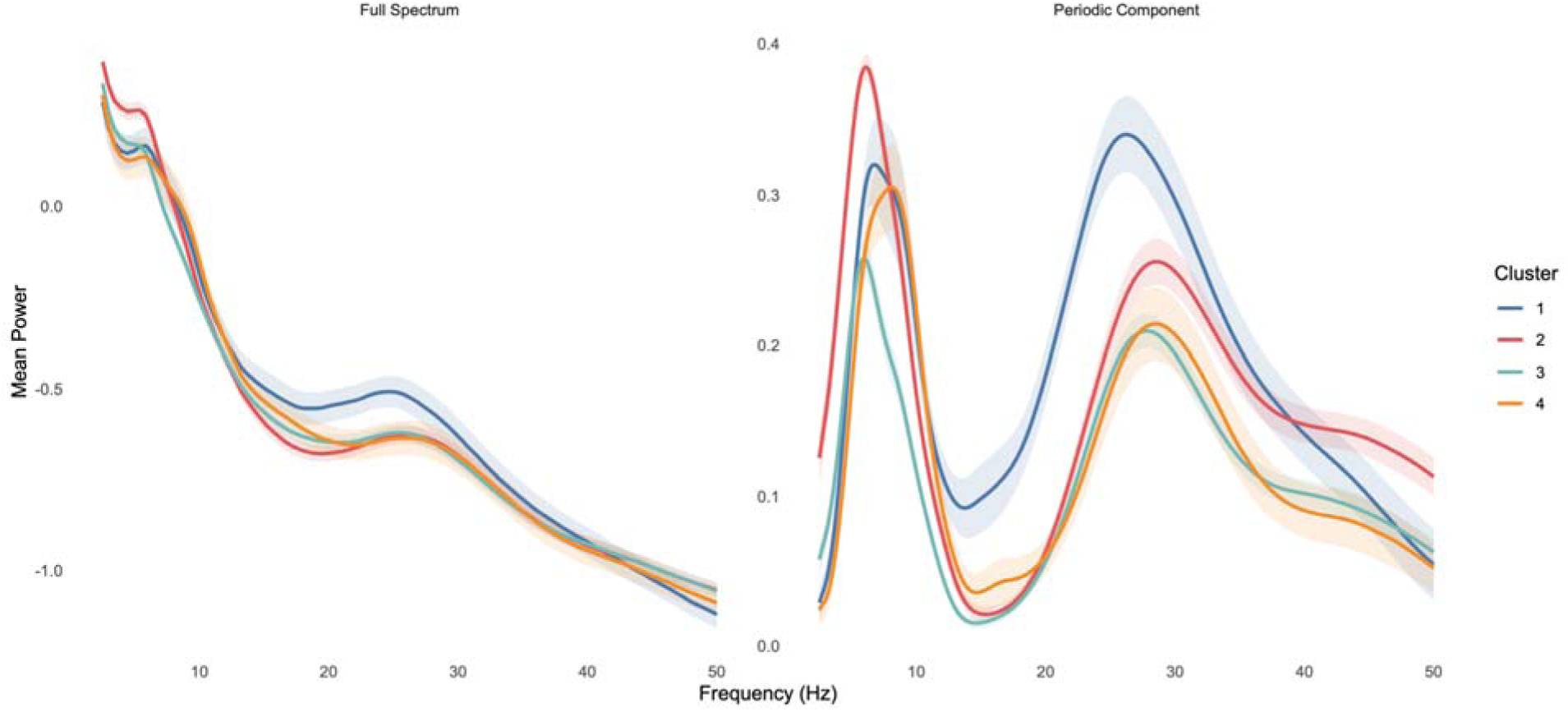
The left panel shows the mean absolute (log-transformed) power spectrum in each cluster, and the right panel shows the mean periodic (aperiodic-adjusted) component in each cluster. These plots are descriptive visualizations of the raw spectral profiles in each cluster and do not reflect the specific features used in clustering, which are presented in Figure 2.

Patterns of EEG features and descriptive statistics in each cluster are reported in Table 3 and visualized in Figure 2. The association between age bin and cluster assignment was not statistically significant, χ²(6, N = 360) = 10.21, *p* = .116, Cramér’s V = .12 (a small effect size), suggesting limited residual age loading after age standardization. Cluster 1 was characterized by both higher alpha power and particularly higher beta power, with moderately higher alpha peak frequency and lower beta peak frequency. Cluster 2 was characterized by particularly higher theta/low alpha power, with moderately lower beta power and moderately lower alpha peak frequency/higher beta peak frequency. Cluster 3 was characterized by broadly lower oscillatory power across alpha and beta bands, alongside lower alpha peak and beta peak frequencies. Cluster 4 was characterized by lower theta power and higher alpha peak frequency, with particularly higher high alpha power and beta peak frequency.

**Figure 2.**
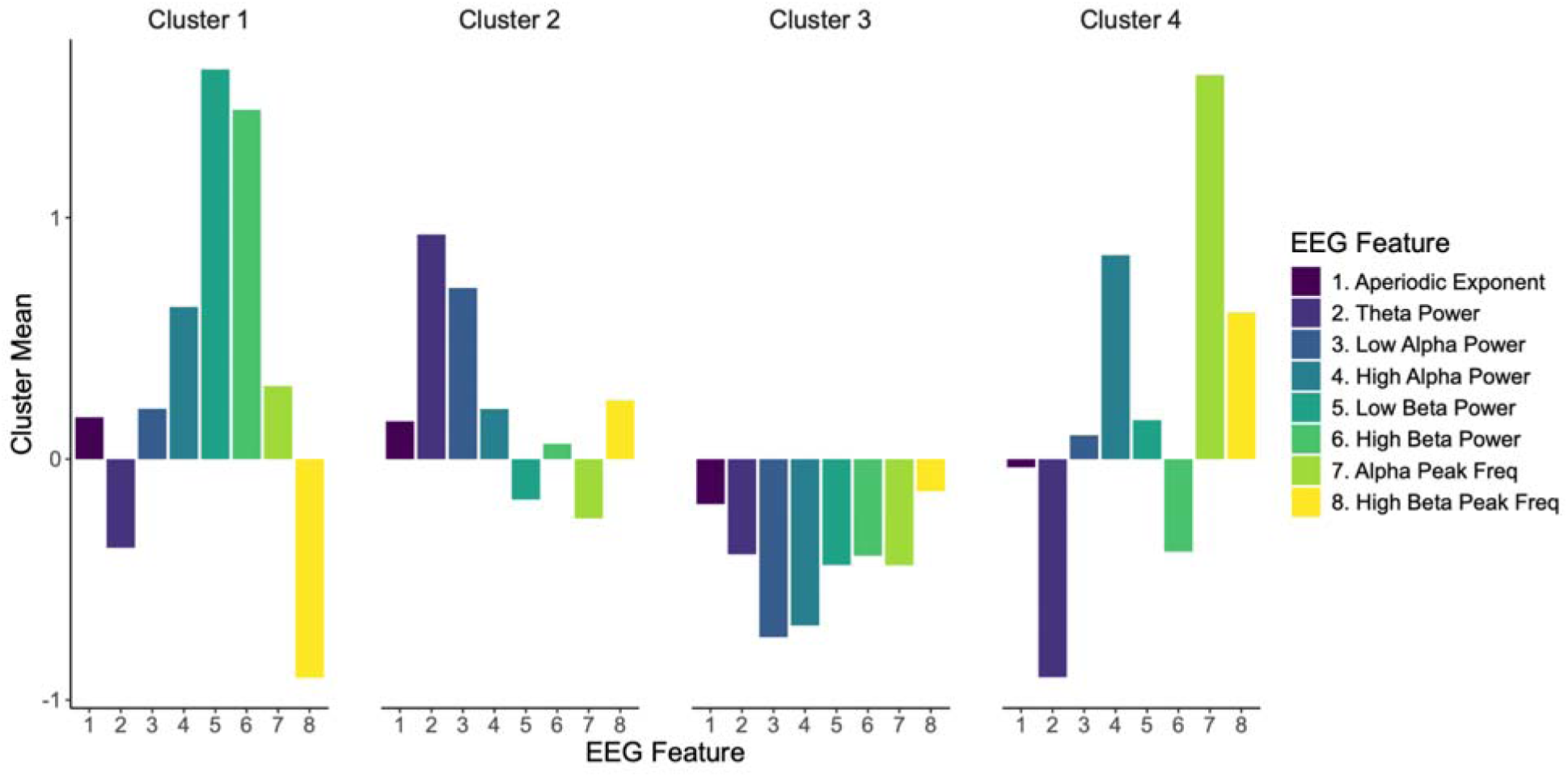
Bar Plots Showing Mean EEG Features in Each Cluster

**Table 3.**
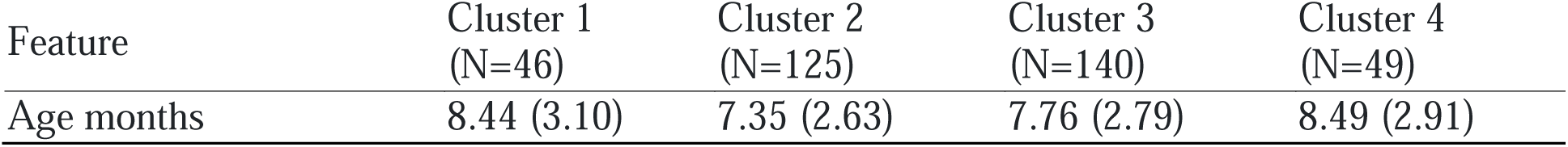
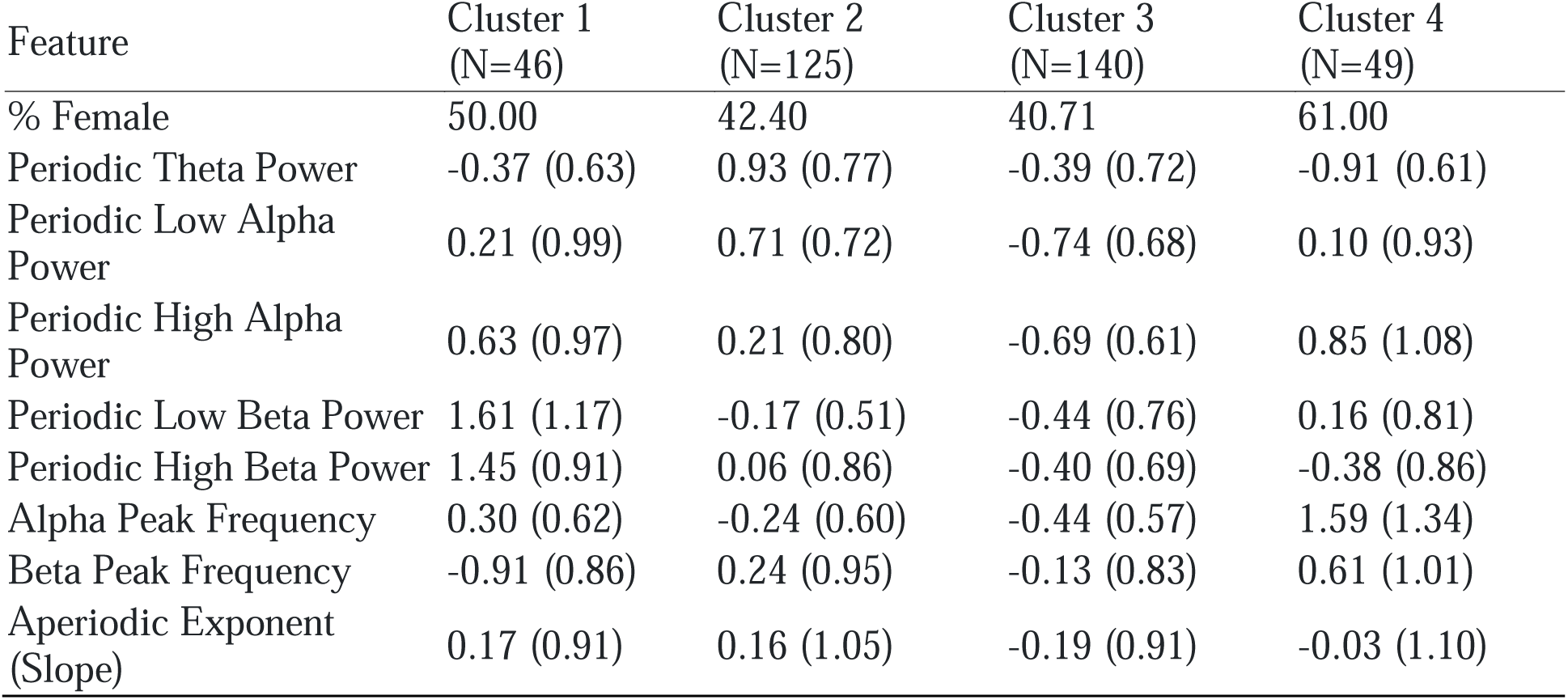
Descriptive statistics for EEG Clusters. Mean (SD)

### Analysis of Cluster Differences in Temperament and Mental Health

To investigate whether infant EEG-derived clusters differentiated temperament and psychopathology through age 7 years, we fit Bayesian mixed-effects models. Analyses included participants with infant EEG cluster assignments who also had follow-up temperament and/or psychopathology data at one or more timepoints (infancy: *n* = 312; age 3: *n* = 204; age 5: *n* = 196; age 7: *n* = 180). Temperament outcomes (surgency, negative affectivity, effortful control [orienting/regulation in infancy]) were assessed longitudinally at infancy and ages 3, 5, and 7 years; psychopathology outcomes (internalizing and externalizing symptoms) were assessed at ages 5 and 7 years. For all five outcomes, additive models including cluster and age provided the best fit relative to cluster-only and cluster-by-age interaction models (Table S2), suggesting limited evidence for systematic age-dependent variation. Accordingly, we focused on age-averaged contrasts for primary inference. However, given established developmental variation across this period, we also examined interaction models, and full within-age contrasts are reported in the supplementary materials. Because the interaction model yields identical marginal (age-averaged) contrasts when age-dependent variation is minimal, while also permitting estimation of age-specific effects, we used the interaction model as the basis for all reported contrasts; age-averaged estimates from this model were closely consistent with those from the additive model. Prior sensitivity analyses demonstrated that the results did not substantially differ according to the prior used.

Posterior probabilities for cluster differences in each domain across ages are shown in Figure 3. Cluster differences were most pronounced for effortful control. Posterior contrasts indicated strong evidence for higher effortful control in Clusters 1 and 2 relative to Clusters 3 and 4 across ages (posterior probabilities > .95), with additional moderate evidence supporting separation between Clusters 1 and 2 (.84), and between Clusters 3 and 4 (0.83). Together, these results support a graded pattern of differentiation across clusters (Cluster 1 > 2 > 3 > 4). Evidence for cluster contrasts across surgency and negative affectivity were more moderate. There was moderate evidence for lower surgency in Cluster 2 relative to Clusters 1, 3, and 4, moderate evidence for lower negative affectivity in Cluster 1 relative to Clusters 2 (.19) and 4 (.10), and moderate-strong evidence for lower negative affectivity in Cluster 3 relative to Clusters 2 (.10) and 4 (.05).

**Figure 3.**
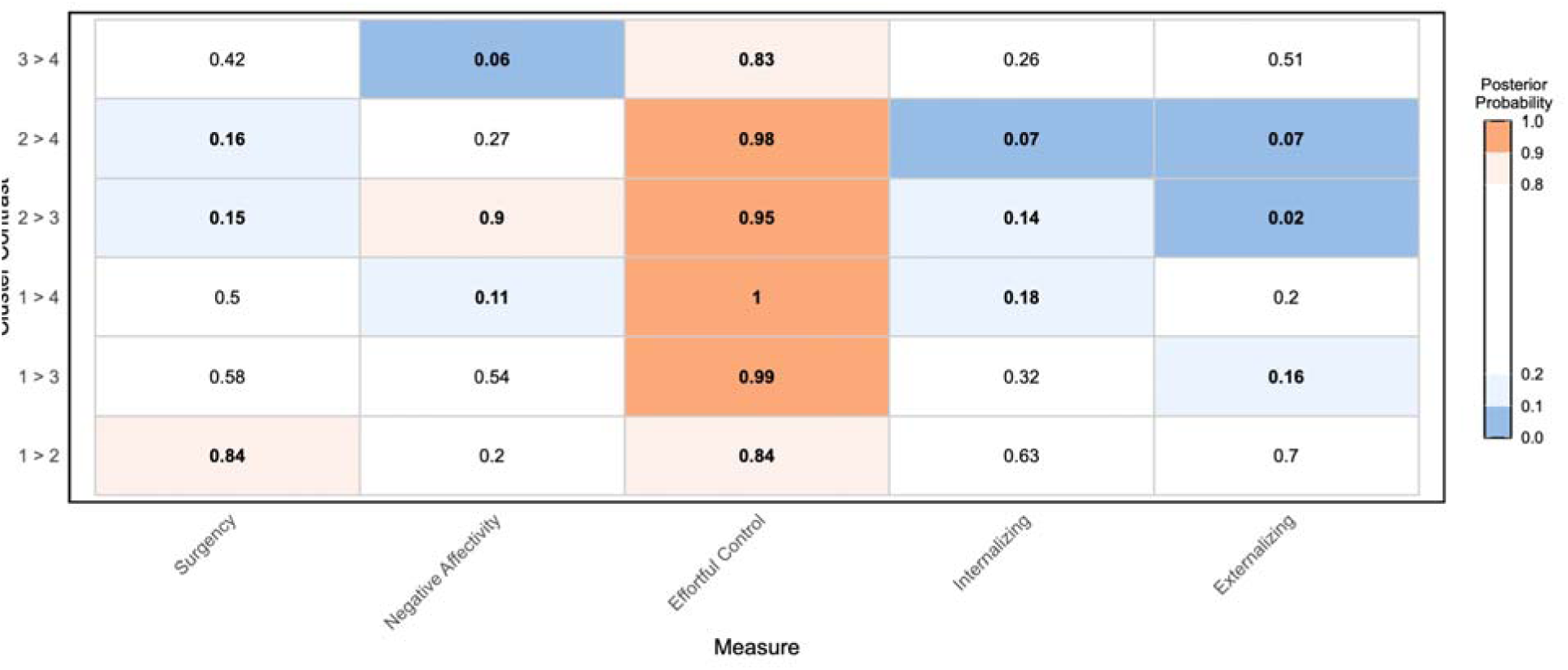
Posterior probabilities of pairwise cluster differences across temperament and psychopathology outcomes. Values represent posterior probabilities that the first cluster in each contrast exhibits higher scores than the second. Temperament outcomes (Surgency, Negative Affectivity, Effortful Control [Orienting/Regulation in infancy]) were assessed in infancy and at ages 3, 5, and 7 years. Psychopathology outcomes (Internalizing and Externalizing) symptoms were assessed at ages 5 and 7 years. Cluster contrasts represent expected marginal means averaged across relevant ages.

For psychopathology, the strongest evidence was observed for lower internalizing symptoms in Cluster 2, which also had higher effortful control and lower surgency, relative to Clusters 3 and 4. There was additional moderate evidence for lower internalizing symptoms in Cluster 1 relative to Cluster 4 (.18) and lower externalizing symptoms in Cluster 1 relative to Cluster 3 (.17). Full posterior contrast estimates, including median Cohen’s d values, and 95% credible intervals for all pairwise cluster comparisons, are provided in Table S3. Within-age contrasts are provided in Figures S5-S6. Consistent with model comparisons favoring additive over interaction models, within-age results broadly aligned with the age-averaged results, with the direction and ordering of cluster differences across timepoints largely preserved, particularly for effortful control. Variability in posterior probabilities across ages was modest to moderate in magnitude and consistent with differences in precision because of sample size as well as potential subtle developmental modulation.

## Discussion

In the current study we applied a data-driven unsupervised clustering pipeline to derive neurophysiological profiles from infant EEG and evaluated how these profiles prospectively differentiated dimensions of temperament and psychopathology in a longitudinal sample with repeated measures from infancy through age 7 years. We applied clustering to infant baseline EEG features, including periodic band power, peak frequencies, and the aperiodic exponent, which yielded four distinct profiles. These profiles robustly differentiated effortful control across childhood, revealing a graded pattern across clusters, with additional differentiation across surgency and negative affectivity. Finally, profiles showed modest differentiation of internalizing and externalizing symptoms, suggesting that these profiles may capture early emerging variation in mental health risk. Taken together, these findings indicate that data-driven neurophysiological profiles derived in infancy prospectively differentiate patterns of temperament and emerging mental health risk, with an organizing gradient along early regulatory capacity.

The four identified neurophysiological profiles were differentiated primarily by distinct configurations of oscillatory power and peak frequencies. Specifically, Cluster 1 was characterized by relatively higher oscillatory power across frequency bands, with particularly high beta, suggesting higher cortical activation and excitability. Cluster 2 also demonstrated relatively strong power, but with a pronounced low frequency dominance, suggestive of a more developmentally typical or balanced spectral organization. In contrast, Cluster 3 was characterized by broadly attenuated power and lower peak frequencies, suggesting lower overall cortical activation and a potentially less mature profile. Finally, Cluster 4 combined relatively low power with higher alpha and beta peak frequencies, indicating a shift toward faster oscillatory dynamics relative to age. These profiles suggest that data-driven clustering of infant EEG captures distinct patterns of neural activity that may provide additional insight into early neural organization beyond individual EEG metrics.

Although these profiles were derived solely from neurophysiological features, they showed a graded pattern of differentiation in effortful control across infancy to age 7 years, with a clear ordering across clusters (Cluster 1>Cluster 2>Cluster 3>Cluster 4) and particularly strong evidence for higher effortful control in Clusters 1 and 2 relative to Clusters 3 and 4. This pattern suggests that infant neurophysiological profiles are organized along a robust axis of effortful control, i.e., the capacity to regulate attention, emotion, and behavior (Rothbart & Bates, 2006). Effortful control is a core developmental construct that underlies the development of emotional and behavioral self-regulation capacities that are important for later mental health outcomes, including internalizing and externalizing psychopathology (Eisenberg et al., 2010). Clusters characterized by relatively strong oscillatory power (Clusters 1 and 2) showed higher effortful control, whereas those with attenuated and frequency-shifted profiles (Clusters 3 and 4) showed lower effortful control. Differences across other temperament domains and early internalizing and externalizing symptoms were more modest but also distinctly patterned, suggesting that effortful control provides a primary axis of differentiation, with additional variation across other temperament domains alongside corresponding downstream differences in early psychopathology symptoms further distinguishing these neurophysiological profiles.

Considered together, the broader pattern of findings further suggests that these profiles may reflect distinct developmental phenotypes. Cluster 2 was characterized by a high-power, low-frequency dominant spectral profile, was associated with higher effortful control and lower surgency, and showed the most consistent pattern of lower risk, with clear directional evidence for lower internalizing and externalizing symptoms relative to Clusters 3 and 4. Cluster 1 also showed relatively high effortful control, but also higher surgency compared to Cluster 2, consistent with its relatively elevated beta power profile, and showed a less robust pattern of lower mental health risk compared to Clusters 3 and 4. In contrast, Clusters 3 and 4 both showed lower effortful control, but with distinct neurophysiological signatures (attenuated power vs. fast-shifted frequency). Notably, Clusters 2 (*N* = 125) and 3 (*N* = 140) comprised larger groups, suggesting that these profiles may reflect more common developmental configurations in this sample, whereas Clusters 1 (*N* = 46) and 4 (*N* = 49) were smaller and may capture more specific risk profiles. There was more limited evidence for clear differentiation between Clusters 1 and 2 and between Clusters 3 and 4 across temperament and psychopathology outcomes. Given that clusters were derived solely from EEG, they may capture neurophysiological variation that does not map directly onto temperament or mental health, or that reflects comparable levels of risk expressed through different underlying mechanisms. Nevertheless, there was some indication of divergence between Clusters 3 and 4, with clearer differentiation in negative affectivity, modest differentiation in effortful control, and directionally consistent but uncertain differences in internalizing symptoms. These profiles may be of particular interest for longitudinal follow up, as they could represent alternative pathways to vulnerability that become more clearly differentiated during later development or differentially interact with environmental risk factors to shape later psychopathology.

The patterns of behavioral differentiation observed across EEG-derived profiles reflect several strengths of the analytic approach used. By applying unsupervised clustering to multiple EEG features, we derived neurophysiological profiles that integrate information across spectral measures while remaining interpretable in terms of the underlying patterns of neural organization they captured. Because these profiles were derived solely from infant EEG, they capture biologically defined patterns of variation in neural function during one of the earliest measurable developmental windows, independent of categorical diagnoses or behavioral measures. The fact that these biologically defined profiles differentiated temperament traits (particularly effortful control) and internalizing and externalizing symptoms, emphasizes the potential utility of such biologically grounded approaches for identifying early neurodevelopmental profiles with relevance to later mental health risk during a developmental period in which symptom presentations typically remain non-specific and transdiagnostic.

To derive neurophysiological profiles, we employed an unsupervised clustering pipeline using EEG features derived from spectral parameterization, with dimensionality reduction, and Bayesian model averaged clustering. We used spectral parameterization to derive principled features capturing both periodic and aperiodic activity. For interpretability and consistent with much infant-oriented work, we opted for whole-scalp averages; future work may extend this framework to incorporate spatially resolved features, as well as incorporate additional EEG metrics (e.g., functional connectivity), or even multimodal biological data. Although the current sample size placed fewer constraints on feature dimensionality relative to smaller studies, we applied PCA to reduce collinearity and stabilize clustering; this approach additionally supports scalability to higher-dimensional feature spaces in future work. The aperiodic exponent loaded strongly on PC4, thus aperiodic variation may have contributed less to cluster differentiation than periodic features. Although clusters were defined by distinct periodic structure, this reduced aperiodic contribution may warrant consideration in future work given prior associations between the aperiodic exponent and temperament in early life (e.g., (Sacks, Levin, et al., 2025)).

We used Bayesian model averaging to integrate three complementary clustering algorithms, reducing dependence on any single algorithm, and assigned a single cluster per individual based on aggregated probabilities. This approach can be readily extended to alternative clustering algorithms or explicit modeling of assignment uncertainty in future work. Given the strong association between infant EEG features and age (An et al., 2025) we standardized features within recruitment age bins to avoid clusters primarily reflecting age differences and instead capture age-relative neurophysiological organization. We used Bayesian mixed-effects models to enable probabilistic inference for between-cluster differences and extend prior cross-sectional work longitudinally. We focused primary interpretations on probabilistic cluster-level contrasts given the study aims and intuitive interpretability of probability measures, with effect sizes and age-specific estimates additionally reported in supplementary materials. This Bayesian framework could also be extended for clinically oriented applications in future work, including approaches such as estimation of individual-level probabilities (i.e., individual risk prediction) and incorporation of decision-relevant thresholds.

Furthermore, the study should be interpreted in the context of broader sample strengths and limitations. The relatively large sample for this age group with repeated measures across childhood represents a major strength. However, the sample comprised predominantly White families of middle to high socioeconomic status within a single U.S. geographic region, with relatively low symptom levels compared to clinical populations. Replication in more diverse and clinically enriched samples will be important to establish the generalizability of both the identified neurophysiological profiles and their associations with temperament and psychopathology. However, the fact that profiles derived solely from infant EEG showed prospective differentiation of temperament, as well as internalizing and externalizing symptoms even within a non-clinical sample is encouraging.

## Conclusions

In this study, we applied data-driven clustering to derive neurophysiological profiles from infant baseline EEG and examined how these profiles prospectively differentiated temperament and psychopathology symptoms across childhood. We identified four neurophysiological profiles, which prospectively differentiated early regulatory capacity and emerging psychopathology symptoms in childhood. EEG-derived profiles separated most clearly along an effortful control axis, with profiles associated with higher effortful control also characterized by lower internalizing and externalizing symptoms, and profiles associated with lower effortful control also showing higher psychopathology symptoms. Together, these findings highlight the potential of data-driven EEG profiling as a scalable, biologically informed approach that, with ongoing study, may support early risk differentiation prior to the consolidation of stable psychiatric diagnoses.

## Data Availability

The data that support the findings of this study are available from the corresponding author upon reasonable request. Code used for the clustering and statistical analyses has been deposited in OSF and will be made publicly available upon publication of the peer-reviewed version of this manuscript.

## Notes

### Competing Interest Statement

The authors have declared no competing interest.

### Author Declarations

The Institutional Review Board at Boston Children's Hospital approved all study methods and procedures.

### Funding

- National Institute of Mental Health: MH078829

## References

Achenbach, T., & Rescorla, L. (2000). Child behavior checklist for ages 1 1/2-5. Reporter, 10, 20.

Achenbach, T. M., Dumenci, L., & Rescorla, L. A. (2001). Ratings of relations between DSM-IV diagnostic categories and items of the CBCL/6-18, TRF, and YSR. Burlington, VT: University of Vermont, 1–9.

Achenbach, T. M., & Edelbrock, C. (1991). Child behavior checklist. Burlington (Vt), 7, 371–392.

Achenbach, T. M., & Rescorla, L. A. (2014). The Achenbach system of empirically based assessment (ASEBA) for ages 1.5 to 18 years. In The use of psychological testing for treatment planning and outcomes assessment (pp. 179–213). Routledge.

An, W. W., Bhowmik, A. C., Nelson, C. A., & Wilkinson, C. L. (2025). EEG-based brain age prediction in infants-toddlers: Implications for early detection of neurodevelopmental disorders. Dev Cogn Neurosci, 71, 101493. 10.1016/j.dcn.2024.101493

Arias, D., Saxena, S., & Verguet, S. (2022). Quantifying the global burden of mental disorders and their economic value. eClinicalMedicine, 54. 10.1016/j.eclinm.2022.101675

Bufferd, S. J., Dougherty, L. R., Carlson, G. A., & Klein, D. N. (2011). Parent-reported mental health in preschoolers: findings using a diagnostic interview. Compr Psychiatry, 52(4), 359–369. 10.1016/j.comppsych.2010.08.006

Bürkner, P.-C. (2017). brms: An R Package for Bayesian Multilevel Models Using Stan. Journal of Statistical Software, 80(1). 10.18637/jss.v080.i01

Chung, H., Job Said, A., Tager-Flusberg, H., Nelson, C. A., & Wilkinson, C. L. (2025). The association between infant EEG aperiodic exponent and the trajectory of restricted and repetitive behaviors for toddlers with and without autism. Journal of Neurodevelopmental Disorders, 17(1), 58. 10.1186/s11689-025-09651-3

Conway, C. C., Forbes, M. K., & South, S. C. (2022). A Hierarchical Taxonomy of Psychopathology (HiTOP) Primer for Mental Health Researchers. Clinical Psychological Science, 10(2), 236–258. 10.1177/21677026211017834

Copeland, W. E., Wolke, D., Shanahan, L., & Costello, E. J. (2015). Adult Functional Outcomes of Common Childhood Psychiatric Problems: A Prospective, Longitudinal Study. JAMA Psychiatry, 72(9), 892–899. 10.1001/jamapsychiatry.2015.0730

Desgraupes, B. (2023). clusterCrit: Clustering Indices. In (Version 1.3.0) https://CRAN.R-project.org/package=clusterCrit

Donoghue, T., Haller, M., Peterson, E. J., Varma, P., Sebastian, P., Gao, R., Noto, T., Lara, A. H., Wallis, J. D., Knight, R. T., Shestyuk, A., & Voytek, B. (2020). Parameterizing neural power spectra into periodic and aperiodic components. Nature Neuroscience, 23(12), 1655–1665. 10.1038/s41593-020-00744-x

Egger, H. L., & Angold, A. (2006). Common emotional and behavioral disorders in preschool children: presentation, nosology, and epidemiology. Journal of Child Psychology and Psychiatry, 47(3-4), 313–337. 10.1111/j.1469-7610.2006.01618.x

Eisenberg, N., Spinrad, T. L., & Eggum, N. D. (2010). Emotion-related self-regulation and its relation to children’s maladjustment. Annu Rev Clin Psychol, 6, 495–525. 10.1146/annurev.clinpsy.121208.131208

Forbes, O., Santos-Fernandez, E., Wu, P. P., Xie, H. B., Schwenn, P. E., Lagopoulos, J., Mills, L., Sacks, D. D., Hermens, D. F., & Mengersen, K. (2023). clusterBMA: Bayesian model averaging for clustering. PLoS One, 18(8), e0288000. 10.1371/journal.pone.0288000

Forbes, O., Schwenn, P. E., Wu, P. P., Santos-Fernandez, E., Xie, H. B., Lagopoulos, J., McLoughlin, L. T., Sacks, D. D., Mengersen, K., & Hermens, D. F. (2022). EEG-based clusters differentiate psychological distress, sleep quality and cognitive function in adolescents. Biol Psychol, 173, 108403. 10.1016/j.biopsycho.2022.108403

Gabard-Durnam, L. J., Mendez Leal, A. S., Wilkinson, C. L., & Levin, A. R. (2018). The Harvard Automated Processing Pipeline for Electroencephalography (HAPPE): Standardized Processing Software for Developmental and High-Artifact Data [Technology Report]. Frontiers in Neuroscience, 12. 10.3389/fnins.2018.00097

Kessler, R. C., Berglund, P., Demler, O., Jin, R., Merikangas, K. R., & Walters, E. E. (2005). Lifetime prevalence and age-of-onset distributions of DSM-IV disorders in the National Comorbidity Survey Replication. Arch Gen Psychiatry, 62(6), 593–602. 10.1001/archpsyc.62.6.593

Lenth, R. V., & Piaskowski, J. (2025). emmeans: Estimated Marginal Means, aka Least-Squares Means. 10.32614/CRAN.package.emmeans

Levin, A. R., Méndez Leal, A. S., Gabard-Durnam, L. J., & O’Leary, H. M. (2018). BEAPP: The Batch Electroencephalography Automated Processing Platform. Frontiers in Neuroscience, 12. 10.3389/fnins.2018.00513

McVoy, M., Lytle, S., Fulchiero, E., Aebi, M. E., Adeleye, O., & Sajatovic, M. (2019). A systematic review of quantitative EEG as a possible biomarker in child psychiatric disorders. Psychiatry Res, 279, 331–344. 10.1016/j.psychres.2019.07.004

Mouselimis, L. (2020). ClusterR: Gaussian Mixture Models, K-Means, Mini-Batch-Kmeans, K-Medoids and Affinity Propagation Clustering. R package version 1.2. 2. In.

Nelson, P. M., Sacks, D. D., Korom, M., Remec, N., Perkins, G. A., Cook, K. M., Stoyell, S. M., Evans, M. E., Moser, J., Capparini, C., Inala, S., Huszar, I. N., Zieff, M. R., Vernetti, A., Norton, E. S., Wilkinson, C. L., & Arichi, T. (2026). Bridging the gap: Translating fetal, infant, and toddler neuroimaging insights into clinical practice. Dev Cogn Neurosci, 79, 101696. 10.1016/j.dcn.2026.101696

R Core Team. (2025). R: A Language and Environment for Statistical Computing. In R Foundation for Statistical Computing. https://www.R-project.org/

Rothbart, M. K., & Bates, J. E. (2006). Temperament. In Handbook of child psychology: Social, emotional, and personality development, *Vol.* 3, 6th *ed.* (pp. 99–166). John Wiley & Sons, Inc.

Sacks, D. D., Levin, A. R., Nelson, C. A., & Bosquet Enlow, M. (2025). Associations Among EEG Aperiodic Slope, Infant Temperament, and Maternal Anxiety/Depression Symptoms in Infancy. Psychophysiology, 62(1), e14757. 10.1111/psyp.14757

Sacks, D. D., Valdes, V., Wilkinson, C. L., Levin, A. R., Nelson, C. A., & Bosquet Enlow, M. (2025). Longitudinal Trajectories of Aperiodic EEG Activity in Early to Middle Childhood. Child Development, 96(5). 10.1111/cdev.14261

Sawrikar, V., Macbeth, A., Gillespie-Smith, K., Brown, M., Lopez-Williams, A., Boulton, K., Guestella, A., & Hickie, I. (2022). Transdiagnostic Clinical Staging for Childhood Mental Health: An Adjunctive Tool for Classifying Internalizing and Externalizing Syndromes that Emerge in Children Aged 5–11 Years. Clinical Child and Family Psychology Review, 25(3), 613–626. 10.1007/s10567-022-00399-z

Vergunst, F., Commisso, M., Geoffroy, M.-C., Temcheff, C., Poirier, M., Park, J., Vitaro, F., Tremblay, R., Côté, S., & Orri, M. (2023). Association of Childhood Externalizing, Internalizing, and Comorbid Symptoms With Long-term Economic and Social Outcomes. JAMA Network Open, 6(1), e2249568–e2249568. 10.1001/jamanetworkopen.2022.49568

Wilkinson, C. L., Yankowitz, L. D., Chao, J. Y., Gutiérrez, R., Rhoades, J. L., Shinnar, S., Purdon, P. L., & Nelson, C. A. (2024). Developmental trajectories of EEG aperiodic and periodic components in children 2–44 months of age. Nature communications, 15(1), 5788. 10.1038/s41467-024-50204-4

Winkler, I., Debener, S., Müller, K. R., & Tangermann, M. (2015, 25-29 Aug. 2015). On the influence of high-pass filtering on ICA-based artifact reduction in EEG-ERP. 2015 37th Annual International Conference of the IEEE Engineering in Medicine and Biology Society (EMBC),

